# Clinical Impact, Costs, and Cost-Effectiveness of Expanded SARS-CoV-2 Testing in Massachusetts

**DOI:** 10.1101/2020.07.23.20160820

**Authors:** Anne M. Neilan, Elena Losina, Audrey C. Bangs, Clare Flanagan, Christopher Panella, G. Ege Eskibozkurt, Amir Mohareb, Emily P. Hyle, Justine A. Scott, Milton C. Weinstein, Mark J. Siedner, Krishna P. Reddy, Guy Harling, Kenneth A. Freedberg, Fatma M. Shebl, Pooyan Kazemian, Andrea L. Ciaranello

**Affiliations:** Division of General Academic Pediatrics, Department of Pediatrics, Massachusetts General Hospital, Boston, MA; Division of Infectious Diseases, Department of Medicine, Massachusetts General Hospital, Boston, MA; Medical Practice Evaluation Center, Massachusetts General Hospital, Boston, MA; Harvard Medical School, Boston, MA; Orthopedic and Arthritis Center for Outcomes Research (OrACORe), Department of Orthopedic Surgery, Brigham and Women’s Hospital, Boston, MA; Policy and Innovation eValuation in Orthopedic Treatments (PIVOT) Center, Department of Orthopedic Surgery, Brigham and Women’s Hospital, Boston, MA; Department of Biostatistics, Boston University School of Public Health, Boston, MA; Harvard University Center for AIDS Research, Cambridge, MA; Department of Health Policy and Management, Harvard T.H. Chan School of Public Health, Boston, MA, USA; Africa Health Research Institute, KwaZulu-Natal, South Africa; Division of Pulmonary and Critical Care Medicine, Department of Medicine, Massachusetts General Hospital, Boston, MA; Department of Epidemiology, Harvard T.H. Chan School of Public Health, Boston, MA; Institute for Global Health, University College London, London, UK; MRC/Wits Rural Public Health & Health Transitions Research Unit (Agincourt), University of the Witwatersrand, Johannesburg, South Africa; Division of General Internal Medicine, Department of Medicine, Massachusetts General Hospital, Boston, MA

**Keywords:** COVID-19, testing, screening, PCR, cost-effective

## Abstract

**Background:** We projected the clinical and economic impact of alternative testing strategies on COVID-19 incidence and mortality in Massachusetts using a microsimulation model.

**Methods:** We compared five testing strategies: 1) PCR-severe-only: PCR testing only patients with severe/critical symptoms; 2) Self-screen: PCR-severe-only plus self-assessment of COVID-19-consistent symptoms with self-isolation if positive; 3) PCR-any-symptom: PCR for any COVID-19-consistent symptoms with self-isolation if positive; 4) PCR-all: PCR-any-symptom and one-time PCR for the entire population; and, 5) PCR-all-repeat: PCR-all with monthly re-testing. We examined effective reproduction numbers (R_e_, 0.9-2.0) at which policy conclusions would change. We used published data on disease progression and mortality, transmission, PCR sensitivity/specificity (70/100%) and costs. Model-projected outcomes included infections, deaths, tests performed, hospital-days, and costs over 180-days, as well as incremental cost-effectiveness ratios (ICERs, $/quality-adjusted life-year [QALY]).

**Results:** In all scenarios, PCR-all-repeat would lead to the best clinical outcomes and PCR-severe-only would lead to the worst; at R_e_ 0.9, PCR-all-repeat vs. PCR-severe-only resulted in a 63% reduction in infections and a 44% reduction in deaths, but required >65-fold more tests/day with 4-fold higher costs. PCR-all-repeat had an ICER <$100,000/QALY only when R_e_ ≥1.8. At all R_e_ values, PCR-any-symptom was cost-saving compared to other strategies.

**Conclusions:** Testing people with any COVID-19-consistent symptoms would be cost-saving compared to restricting testing to only those with symptoms severe enough to warrant hospital care. Expanding PCR testing to asymptomatic people would decrease infections, deaths, and hospitalizations. Universal screening would be cost-effective when paired with monthly retesting in settings where the COVID-19 pandemic is surging.

## INTRODUCTION

Local and national testing strategies during the COVID-19 pandemic have varied widely based on geography, supply chain limitations, and political considerations. Countries such as Iceland and South Korea initiated early, widespread testing campaigns targeting people with and without symptoms [1,2]. In the United States, restricted testing capacity early in the pandemic led states such as Massachusetts to test only severely symptomatic people and/or those with known exposure [3]. Despite the variable clinical sensitivity of SARS-CoV-2 polymerase chain reaction (PCR) testing, expanded testing programs could reduce transmissions by increasing isolation of infectious people, thereby reducing hospitalizations and deaths. Testing programs could also allow for the safer resumption of economic and social activity, by providing surveillance for any “second wave” of infection [4].

Massachusetts experienced a major COVID-19 outbreak beginning in March 2020 after a biotechnology convention, which was subsequently fueled by transmission in communities living in multi-generational and multi-family housing [5]. Since new infections peaked in late April [6], Massachusetts has used test positivity rates as a key indicator to guide gradual re-opening, after implementing strategies to reduce transmission risk [4]. In Massachusetts and elsewhere, planning is essential for utilization of key limited resources, such as testing and hospital beds. Our goal was to examine the clinical and economic impact of screening strategies on COVID-19 in Massachusetts.

## METHODS

### Analytic overview

We developed a dynamic state-transition microsimulation model, the CEACOV (Clinical and Economic Analysis of COVID-19 Interventions) model, to reflect the natural history, diagnosis, and treatment of COVID-19. We modeled five testing strategies for all Massachusetts residents (excluding those residing in long-term care facilities): 1) PCR-severe-only: PCR testing only of those who develop severe illness (*i.e*., warranting hospital care), reflecting common practices in Massachusetts through late April 2020 [3]; 2) Self-screen: PCR-severe-only and individuals self-assess the presence of COVID-19-consistent symptoms, using available smartphone applications or websites, and self-isolate if positive [7]; 3) PCR-any-symptom: PCR-severe-only and PCR for people with any COVID-19-consistent symptoms who self-isolate if positive; 4) PCR-all: PCR-any-symptom and a one-time PCR for the entire population; 5) PCR-all-repeat: PCR-all and re-testing every 30 days of those who test negative and remain asymptomatic (Supplementary Figure 1). For those who are not hospitalized, we assume a positive PCR test leads to community self-isolation and is more effective than symptom-based self-isolation. We projected clinical outcomes (infections, COVID-19-related mortality, quality-adjusted life-years [QALY]), and COVID-19-related resource utilization (tests, hospital and intensive care unit (ICU) beds, self-isolation days), and costs for Massachusetts (6.9 million people, excluding long-term care facility residents) over a 180-day horizon. We report incremental cost-effectiveness ratios (ICER: difference in cost divided by difference in quality-adjusted life-years [$/QALY]) from a healthcare sector perspective (Supplementary Methods). The threshold at which interventions are considered cost-effective is a normative value that varies by setting; for the sake of interpretability, we define a strategy as “cost-effective” if its ICER is below $100,000/QALY [8].

### CEACOV model structure

#### Cohort and disease progression

At model start, a closed pre-intervention cohort is seeded with a user-defined proportion of age-stratified individuals (0-19, 25-59, ≥60 years) who are infected with or are susceptible to the SARS-CoV-2 virus. If infected, individuals face daily age-stratified probabilities of disease progression through seven health/disease states, including latent infection, asymptomatic illness, mild/moderate illness, severe illness (warranting hospitalization), critical illness (warranting intensive care), recuperation, and recovery (Supplementary Figure 2). We assume recovered individuals are immune from repeat infection for the 180-day modeled horizon [9]. Susceptible and recovered individuals may also present for testing with symptoms due to non-COVID-19 conditions (“COVID-19-like illness”).

#### Testing

Individuals may experience a daily probability of undergoing SARS-CoV-2 testing. Each PCR testing strategy includes test sensitivity/specificity, turnaround time, and testing frequency.

#### Transmission

In the model, infected individuals have an equal probability of contacting susceptible individuals and transmitting SARS-CoV-2. The effective reproduction number (R_e_) captures the average number of secondary cases per infected individual in the cohort; based on Massachusetts data, this was estimated to be 0.9 in late April 2020 (Supplementary Methods and Supplementary Table 1). People with a positive test result or symptom screen can isolate in the community or in the hospital, which further decreases transmission.

**Table 1.** Input parameters for a model of COVID-19 disease and testing in Massachusetts

| Parameter | Value |  |
| --- | --- | --- |
| **Cohort characteristics** |  |  |
| SARS-CoV-2 prevalence on May 1, 2020, % <sup>a</sup> | 2.99 |  |
| Initial age distribution of cohort, % [10] |  |  |
| 0-19 years | 25 |  |
| 20-59 | 56 |  |
| ≥60 | 19 |  |
| Initial distribution of health states on May 1, 2020, % [11] <sup>a</sup> |  |  |
| Susceptible | 89.38 |  |
| Latent | 0.52 |  |
| Asymptomatic | 0.91 |  |
| Mild/moderate illness | 1.49 |  |
| Severe illness | 0.04 |  |
| Critical illness | 0.02 |  |
| Recuperation | 0.01 |  |
| Recovered | 7.63 |  |
| Health state transition probabilities, by ultimate stage of disease, daily [12–14,16] <sup>b</sup> |  |  |
| Asymptomatic |  |  |
| Latent to asymptomatic | 0.323 |  |
| Asymptomatic to recovered | 0.099 |  |
| Mild/moderate |  |  |
| Latent to asymptomatic | 0.323 |  |
| Asymptomatic to mild/moderate | 0.394 |  |
| Mild/moderate to recovered | 0.095 |  |
| Severe | With hospital care | Without hospital care |
| Latent to asymptomatic | NA | 0.323 |
| Asymptomatic to mild/moderate | NA | 0.394 |
| Mild/moderate to severe | NA | 0.143 |
| Severe to recovered | 0.091 | 0.063 |
| Critical |  |  |
| Latent to asymptomatic | NA | 0.323 |
| Asymptomatic to mild/moderate | NA | 0.394 |
| Mild/moderate to severe | NA | 0.284 |
| Severe to recovered | 0.026 | 0.000 |
| Severe to critical | 0.105 | 0.143 |
| Critical to recuperation | 0.049 | 0.000 |
| Recuperation to recovered | 0.161 | 0.000 |

**Table 1.** Input parameters for a model of COVID-19 disease and testing in Massachusetts (continued)
| Parameter | Value |  |
| --- | --- | --- |
| COVID-19-related mortality while critically ill, probability, daily [42] | With hospital care | Without hospital care |
| 0-19 years | 0.00001 | 0.118 |
| 20-59 | 0.004 | 0.166 |
| ≥60 | 0.050 | 0.203 |
| Development of COVID-19-like illness symptoms among susceptible and recovered, probability, daily [42] |  |  |
| Mild/moderate illness |  |  |
| 0-19 years | 0.00005 |  |
| 20-59 | 0.00005 |  |
| ≥60 | 0.00008 |  |
| Severe illness |  |  |
| 0-19 years | 0.00032 |  |
| 20-59 | 0.00036 |  |
| ≥60 | 0.00053 |  |
| Critical illness |  |  |
| 0-19 years | 0.00009 |  |
| 20-59 | 0.00010 |  |
| ≥60 | 0.00015 |  |
| Presentation to hospital care with severe symptoms, probability <sup>c</sup> | 0.80 |  |
| **Test characteristics** |  |  |
| Self-screen |  |  |
| Positive result, probability | 0.80 |  |
| PCR test [17,18] |  |  |
| Sensitivity, % <sup>d</sup> | 70 |  |
| Specificity, % | 100 |  |
| Turnaround time, days | 1 |  |
| Test acceptance, probability |  |  |
| Asymptomatic/mild illness/moderate illness | 0.80 |  |
| Critical/severe illness | 1.00 |  |

**Table 1.** Input parameters for a model of COVID-19 disease and testing in Massachusetts (continued)
| Parameter | Value |  |
| --- | --- | --- |
| **Transmissions** |  |  |
| $R_e$ | | |
| May 1 – May 30 | 0.9000 |  |
| By health state, probability, daily [30,43,44] <sup>d</sup> |  |  |
| Latent | 0.0000 |  |
| Asymptomatic | 0.2394 |  |
| Mild/moderate illness | 0.1948 |  |
| Severe illness | 0.0135 |  |
| Critical illness | 0.0107 |  |
| Recuperation | 0.0135 |  |
| Recovery | 0.0000 |  |
| Transmission reduction after test result, % <sup>f</sup> | Screen positive | Screen negative |
| Self-screen |  |  |
| Asymptomatic | 0 | 0 |
| Mild/moderate illness | 20 | N/A |
| PCR-based strategies |  |  |
| Asymptomatic | 65 | 0 |
| Mild/moderate illness | 65 | 0 |
| Severe/critical/recuperation <sup>f</sup> | 90 | 90 |
| **Costs (USD 2020)** |  |  |
| SARS-CoV-2 PCR assay [20] | 51 |  |
| Hospital bed, daily [21–23] | 1,640 |  |
| Intensive care unit, daily [21–23] | 2,680 |  |

Abbreviations: PCR, polymerase chain reaction; $R_e$ , Effective reproduction number; USD, United States dollars
<sup>a</sup> Prevalence and distributions were derived from model validation and calibration as described in the Supplementary Material.
<sup>b</sup> Average days spent in each health state stratified by clinical disease progression severity are presented in Supplementary Table 1. Health state transitions are shown in Supplementary Figure 2.
<sup>c</sup> Assumption; includes those with COVID-19 disease and those with COVID-19-like illness.
<sup>d</sup> Test sensitivity is 0% in the latent phase and otherwise does not vary by disease states.
<sup>e</sup> Daily transmission rates contribute to $R_e$ .
<sup>f</sup> Assumptions for transmission reductions following test result are detailed in the Supplementary Material. In severe/critical/recuperation states, transmission reduction is due to hospitalization and thus is applied to all patients regardless of test result.

#### Resource use

The model tallies tests, COVID-19-related use of hospital and ICU bed-days, as well as days spent self-isolating.

### Model inputs

#### Cohort and disease progression

We derived the initial distribution of COVID-19 disease severity by age from the Massachusetts Census and Department of Public Health (Table 1) [10,11]. Disease progression and COVID-19-related mortality are derived from data from China and Massachusetts and calibrated to deaths in Massachusetts (excluding those occurring long-term care facilities) from mid-March to May 1, 2020 (Table 1 and Supplementary Table 1) [11–16].

#### Testing and associated transmission reduction

PCR test sensitivity/specificity are assumed to be 70%/100% (Table 1) [17,18]. In all strategies, patients with severe or critical illness are eligible for diagnostic testing and are hospitalized regardless of PCR test result. Transmission is reduced by 90% for hospitalized people due to infection control and isolation practices (Table 1 and Supplementary Methods). In Self-screen, self-screening is assumed to detect 80% of COVID-associated symptoms. A positive self-screen is assumed to lead to a 20% reduction in the risk of transmission due to partial self-isolation. In the expanded PCR-based strategies, self-isolation among those in the community with a positive PCR test is more effective than after a symptom-based self-screen (65% transmission reduction, regardless of symptoms) [19]; those who test negative do not self-isolate (incorporating the potential for transmissions associated with false-negative tests). PCR test acceptance is assumed to be 80% for those who are asymptomatic or have mild/moderate illness at the time of testing, and 100% for those with severe or critical illness.

#### Epidemic scenarios

For the first month of the simulation, corresponding to May 1, 2020 to May 31, 2020, R_e_ remains 0.9 (Supplementary Table 1). To account for the uncertain trajectory of the epidemic as reopening plans are implemented, we model three scenarios representing epidemics with distinct R_e_ values, in the absence of expanded testing (*i.e*., PCR-severe-only), beginning on June 1, 2020: 1) Slowing (June 1, 2020 R_e_=0.9), suggesting epidemic growth would remain the same as during May (*e.g*. stay-at-home advisory and non-essential business closures); 2) Intermediate (June 1, 2020 R_e_=1.3), suggesting modest increase in epidemic growth; and, 3) Surging (June 1, 2020 R_e_=2.0), suggesting an R_e_ closer to late March/early April Massachusetts estimates (R_e_=2.5-5.9, Supplementary Table 1). We also identified threshold values for the R_e_ at which policy conclusions would change. Transmission probabilities are based on time spent in health state (Table 1).

#### Costs and cost-effectiveness

PCR test cost is $51 [20]. Patients requiring hospitalization accrue per-day costs (hospital: $1,640; ICU: $2,680) [21–23]. We use projected deaths to estimate quality-adjusted life-years lost per strategy (Supplementary Methods) [24].

### Sensitivity and scenario analyses

In each of the three epidemic growth scenarios, we vary PCR sensitivity (30-100%), test acceptance (15-100% for asymptomatic or mild/moderate symptoms), transmission reduction after a positive test (self-screen: 10-40%; PCR: 33-100%), presentation to hospital with severe disease (50-100%), ICU survival (20-80%), testing program costs (including additional outreach costs of offering PCR testing even if declined, $3-$26), and hospital care costs ($820-$3,880). In multiway sensitivity analyses, we vary key parameters simultaneously. In additional analyses, we examined implementation of these testing strategies on April 1, 2020 vs. May 1, 2020; the R_e_ threshold at which conclusions about the preferred strategy shifted (R_e_ 1.3-2.0); the frequency of retesting in PCR-all-repeat (every 7-30 days); patterns of presenting with COVID-19-like illness; varying estimates of life-years lost due to COVID-19-related mortality; and, the impact of costs associated with lost productivity and averted mortality. Further details of methods, as well as model calibration and validation, are in the Supplementary Material.

## RESULTS

### Base case outcomes

#### Clinical outcomes

All the expanded screening strategies would reduce infections and deaths compared to PCR-severe-only. In all scenarios, PCR-all-repeat would lead to the most favorable clinical outcomes and PCR-severe-only would lead to the least favorable outcomes; in the slowing scenario PCR-all-repeat vs. PCR-severe-only resulted in 210,200 vs. 565,300 infections (63% reduction) and 1,800 vs. 3,200 deaths (44% reduction) (Table 2, top section). As R_e_ increases, compared to PCR-severe-only, more expansive screening strategies would lead to greater reductions in infections and deaths (Table 2, bottom section). As R_e_ increases, the expanded screening strategies, compared with PCR-severe-only, would result in a greater reduction in prevalence and lower reduction in the susceptible proportion of the population (Figures 1A-C).

**Table 2.** Clinical and cost-effectiveness outcomes for a model of COVID-19 disease and testing in Massachusetts

|  | Undiscounted | Undiscounted | Discounted | Undiscounted | Discounted |
| --- | --- | --- | --- | --- | --- |
| | Incident infections,<br>No. <sup>a</sup> | Deaths,<br>No. <sup>a</sup> | Total life-years lost,<br>No. <sup>b</sup> | Healthcare costs,<br>\$ <sup>a</sup> | ICER,<br>\$/QALY <sup>c</sup> |
| **Slowing scenario (June 1, 2020 R<sub>e</sub> 0.9)** |  |  |  |  |  |
| PCR-any-symptom | 316,300 | 2,300 | 7,300 | 393,037,000 | - |
| Self-screen | 422,200 | 2,600 | 8,200 | 428,461,000 | dominated |
| PCR-severe-only | 565,300 | 3,200 | 10,100 | 492,552,000 | dominated |
| PCR-all | 281,000 | 2,100 | 6,700 | 654,741,000 | 394,000 |
| PCR-all-repeat | 210,200 | 1,800 | 5,800 | 2,071,400,000 | 1,540,000 |
| **Intermediate scenario (June 1, 2020 R<sub>e</sub> 1.3)** |  |  |  |  |  |
| PCR-any-symptom | 604,600 | 3,500 | 11,100 | 506,489,000 | - |
| Self-screen | 975,200 | 4,400 | 14,100 | 636,392,000 | dominated |
| PCR-all | 543,900 | 3,000 | 9,700 | 768,358,000 | 181,000 |
| PCR-severe-only | 1,471,100 | 6,300 | 20,100 | 832,028,000 | dominated |
| PCR-all-repeat | 298,300 | 2,100 | 6,800 | 2,111,387,000 | 468,000 |
| **Surging scenario (June 1, 2020 R<sub>e</sub> 2.0)** |  |  |  |  |  |
| PCR-any-symptom | 2,924,200 | 11,800 | 37,600 | 1,421,427,000 | - |
| PCR-all | 2,799,400 | 11,300 | 36,000 | 1,673,911,000 | dominated |
| Self-screen | 3,666,900 | 14,700 | 46,500 | 1,753,092,000 | dominated |
| PCR-severe-only | 4,193,800 | 17,300 | 55,000 | 2,010,507,000 | dominated |
| PCR-all-repeat | 1,232,500 | 5,200 | 16,600 | 2,532,432,000 | 53,000 |

Abbreviations: No., Number; PCR, Polymerase chain reaction; R<sub>e</sub>, Effective reproduction number; \$, US dollars; ICER, incremental cost-effectiveness ratio; QALY, quality-adjusted life-year
<sup>a</sup> Includes 180-day horizon between simulated days May 1, 2020 and November 1, 2020.
<sup>b</sup> Total life-years lost were estimated from COVID-related deaths occurring over 180-days. Details are in the Supplementary Material.
<sup>c</sup> Incremental cost effectiveness ratios are calculated by dividing the difference in total life-years lost by the difference in total healthcare-related costs compared to the next most expensive strategy. Dominated strategies are either more expensive and less effective than another strategy (strong dominance) or a combination of two other strategies (weak dominance). Strategies are listed in order of increasing cost as per cost-effectiveness analysis convention. Total life-years lost are discounted at 3%/year; because all healthcare costs occur in year one, costs are not discounted in the base case. Additional details of calculating ICERs may be found in the Supplementary Material.
Infections, deaths, and life-years lost are rounded to the nearest 100. Costs and ICERs are rounded to the nearest 1,000. In-text results describing percentages are calculated from unrounded results.

**Figure 1.**
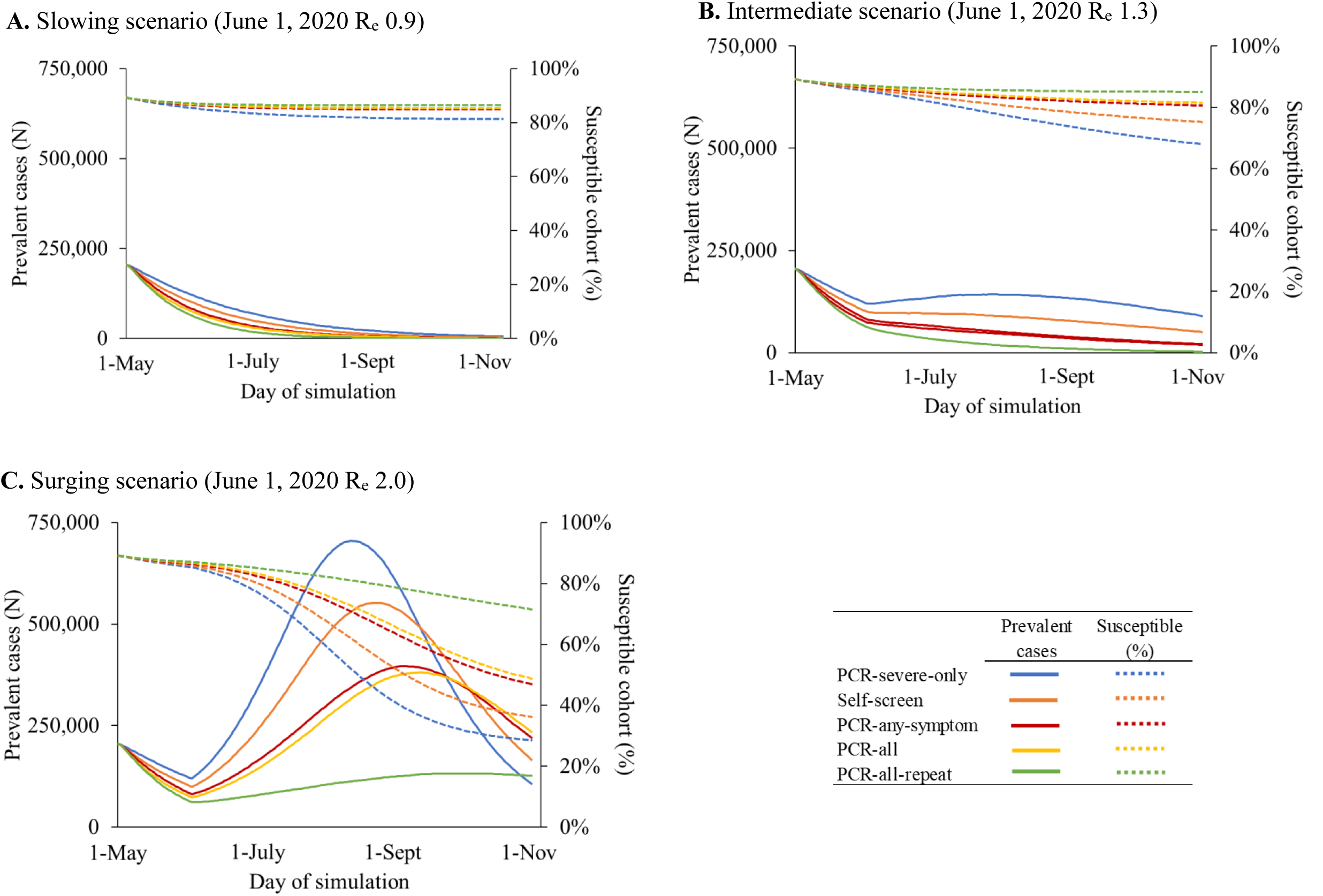
Model-projected SARS-CoV-2 infection prevalence and proportion of susceptible cohort. For the modeled strategies, prevalent COVID-19 cases over time are plotted as solid lines on the left vertical axis, while the percentages of the cohort remaining susceptible to infection over time are plotted as dotted lines on the right vertical axis. People with SARS-CoV-2 are no longer considered prevalent when they have recovered (Supplementary Figure 1). Results shown represent the population of Massachusetts. Testing strategies are denoted by different colored lines. Panel A represents a slowing scenario in which the effective reproduction number (R_e_) on June 1, 2020 is 0.9. Panel B represents an intermediate scenario in which R_e_ one June 1, 2020 is 1.3, and panel C represents a surging scenario in which R_e_ on June 1, 2020 is 2.0. Abbreviations: R_e_, effective reproduction number; PCR, Polymerase chain reaction

#### Resource utilization and costs

In all epidemic growth scenarios, PCR-any-symptoms would lead to lower total costs compared to PCR-severe only. In the slowing scenario, PCR-all-repeat would lead to the greatest reduction in cumulative bed-days compared to PCR-severe-only: 88,500 vs. 139,100 hospital bed-days (36% reduction) and 55,400 vs. 88,500 ICU bed-days (37% reduction) but would require >65-fold times more tests/day (192,500 vs. 2,900) at 4-fold higher total costs ($2.1 billion vs. $493 million) (Tables 2 and 3). In the slowing and intermediate scenarios, peak hospital bed use is similar across all strategies. In the surging scenario, however, all of the other PCR-based strategies would reduce peak hospital and ICU bed use compared to PCR-severe-only: hospital beds (6,200 vs. 2,600-3,600) and ICU beds (3,600 vs. 1,200-2,100) (Table 3, bottom section). Supplementary Table 2 reports results/million people.

**Table 3.** Clinical and resource utilization outcomes for a model of COVID-19 disease and testing in Massachusetts

|  | PCR tests | PCR tests,<br>total | Hospital bed-days |  | ICU bed-days |  | Cumulative self-isolation days |
| --- | --- | --- | --- | --- | --- | --- | --- |
|  | per<br>simulation<br>day, mean |  | Cumulative | Peak | Cumulative | Peak |  |
| Slowing scenario (June 1, 2020 R<sub>e</sub> 0.9) |  |  |  |  |  |  |  |
| PCR-severe-only | 2,900 | 521,300 | 139,100 | 2,700 | 88,500 | 1,300 | - |
| Self-screen | 2,900 | 520,800 | 122,700 | 2,800 | 74,700 | 1,200 | 3,689,700 |
| PCR-any-symptom | 4,900 | 877,900 | 105,000 | 2,800 | 65,500 | 1,200 | 1,836,400 |
| PCR-all | 35,200 | 6,337,500 | 100,200 | 2,800 | 61,500 | 1,200 | 2,012,900 |
| PCR-all-repeat | 192,500 | 34,643,400 | 88,500 | 2,600 | 55,400 | 1,200 | 2,378,500 |
| Intermediate scenario (June 1, 2020 R<sub>e</sub> 1.3) |  |  |  |  |  |  |  |
| PCR-severe-only | 2,900 | 529,800 | 247,800 | 2,700 | 148,400 | 1,300 | - |
| Self-screen | 2,900 | 524,900 | 185,900 | 2,800 | 113,500 | 1,200 | 6,285,300 |
| PCR-any-symptom | 5,700 | 1,032,100 | 138,500 | 2,800 | 84,300 | 1,200 | 2,717,000 |
| PCR-all | 36,200 | 6,522,700 | 130,400 | 2,800 | 81,900 | 1,200 | 2,843,800 |
| PCR-all-repeat | 193,400 | 34,811,600 | 98,600 | 2,600 | 60,900 | 1,200 | 2,895,100 |
| Surging scenario (June 1, 2020 R<sub>e</sub> 2.0) |  |  |  |  |  |  |  |
| PCR-severe-only | 3,100 | 551,000 | 612,400 | 6,200 | 364,300 | 3,600 | - |
| Self-screen | 3,000 | 546,100 | 539,100 | 5,000 | 313,200 | 3,000 | 19,510,600 |
| PCR-any-symptom | 12,600 | 2,267,100 | 408,300 | 3,600 | 236,700 | 2,100 | 9,600,800 |
| PCR-all | 45,000 | 8,094,300 | 397,000 | 3,500 | 226,300 | 2,000 | 9,713,600 |
| PCR-all-repeat | 203,300 | 36,591,000 | 204,600 | 2,600 | 119,000 | 1,200 | 7,674,100 |

Abbreviations: PCR, Polymerase chain reaction; ICU, Intensive care unit; $R_e$ , Effective reproduction number
Includes events occurring during the 180-day horizon between simulated days May 1, 2020 and November 1, 2020. PCR tests, hospital bed-days, ICU bed-days, and self-isolation days are rounded to the nearest 100. In-text results describing percentages are calculated from unrounded results. Cumulative self-isolation days are estimated in addition to the PCR-severe-only strategy.

#### Cost-effectiveness outcomes

Under all epidemic growth scenarios considered, PCR-any-symptom would be clinically superior and cost-saving compared to PCR-severe-only (Table 2). PCR-all-repeat would have an ICER <$100,000/QALY compared to PCR-any-symptom only in the surging scenario ($53,000/QALY). ICERs increase steeply as R_e_ declines (Table 2).

### Sensitivity and scenario analyses

#### Clinical outcomes and resource use

The impact of variation in clinical model input parameters on infections and deaths would be greatest in the surging scenario (Supplementary Figures 3A-F). Varying rates of presentation to hospital care and ICU survival would lead to large changes in mortality, which remain substantial (slowing scenario: 1,400-2,300 deaths/180-days) even under optimistic assumptions (*i.e*., 100% presentation to hospital with severe illness or 80% ICU survival) (Supplementary Figures 3D-F). If expanded PCR testing started April 1, 2020, compared to May 1, 2020, project that PCR-based strategies would have averted 106,200-184,200 infections (Figures 2A-C) and 80-120 deaths in April alone (2D-F).

**Figure 2.**
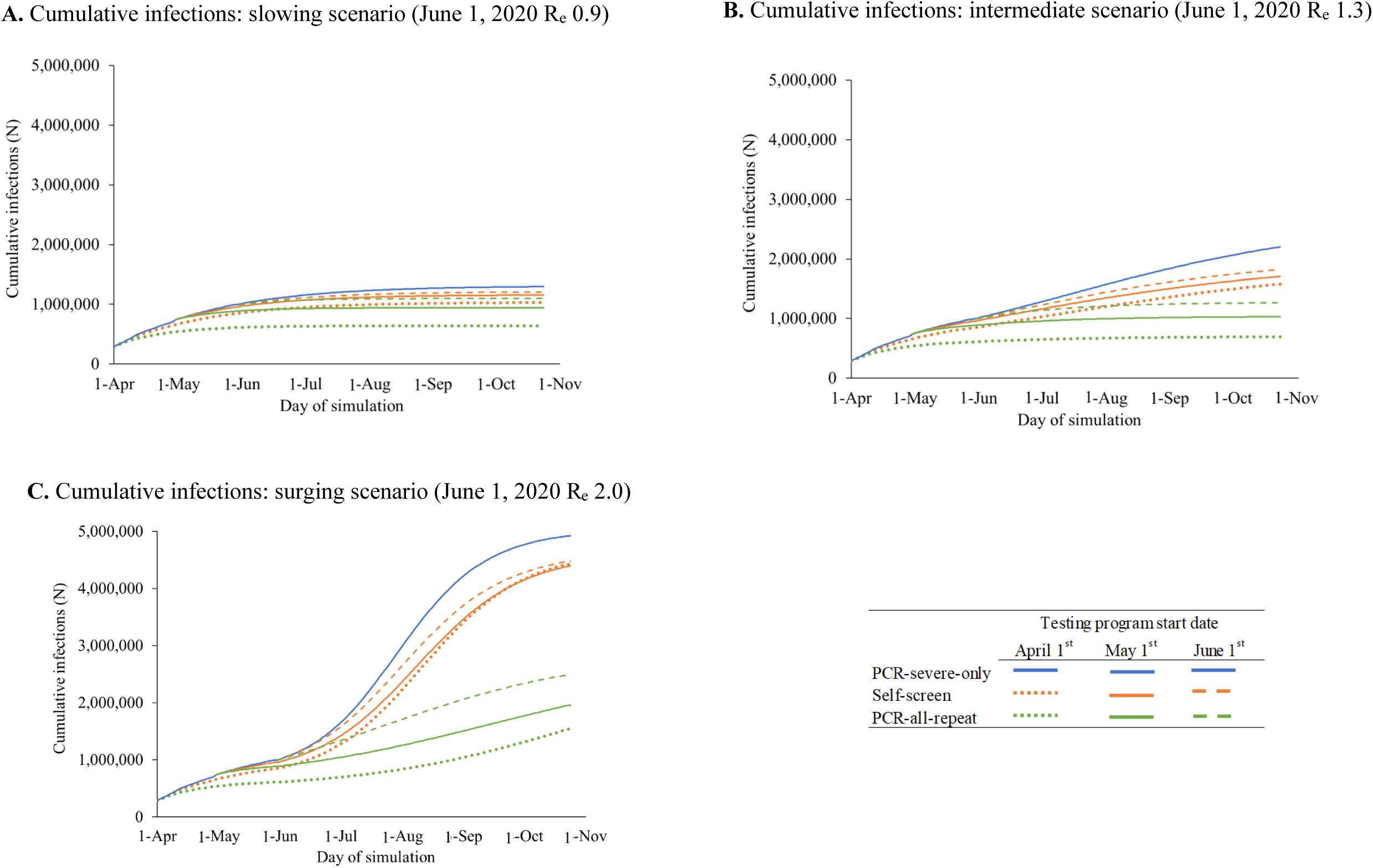
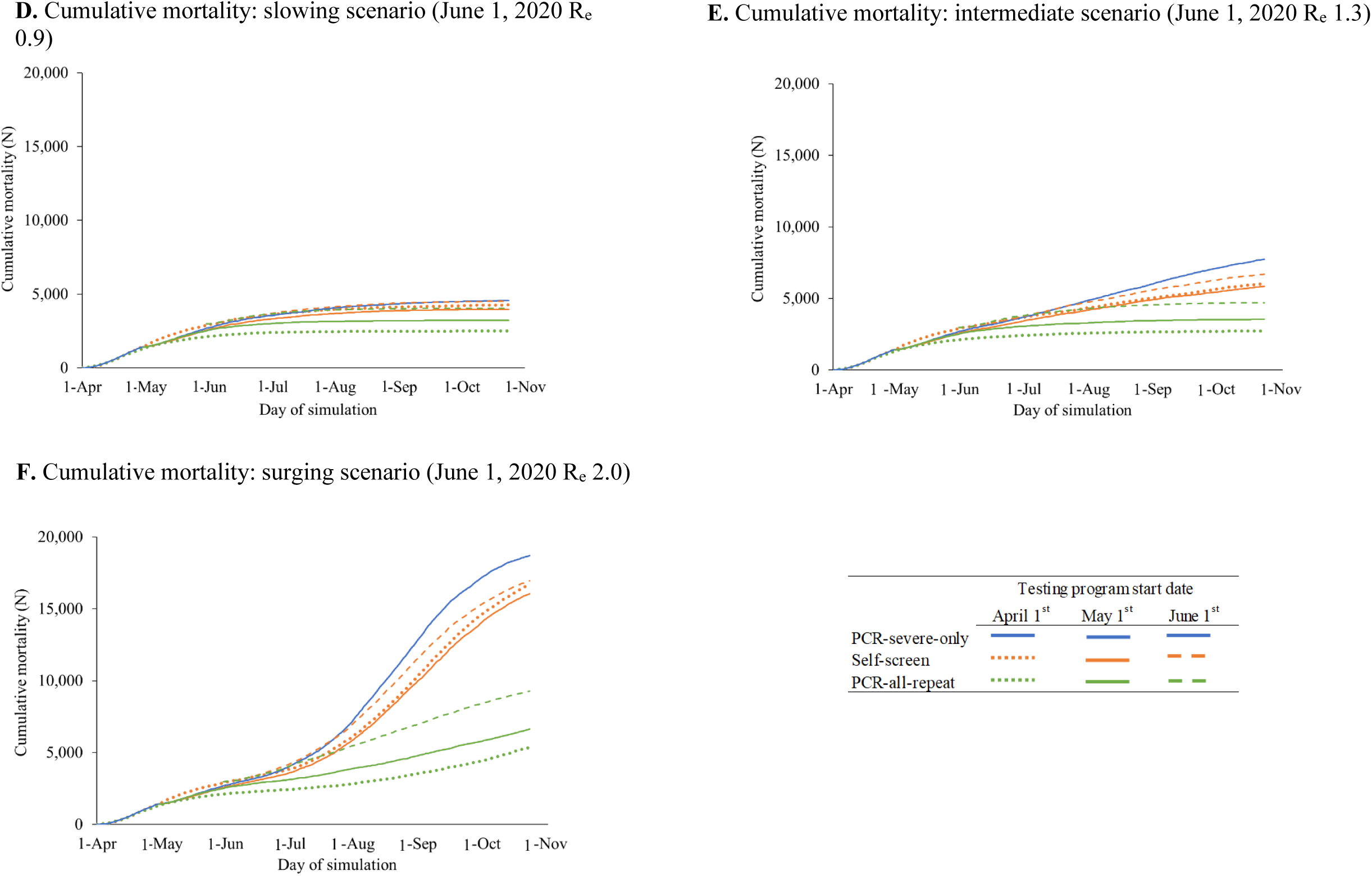
Scenario analyses: Cumulative SARS-CoV-2 infections and mortality resulting from alternate dates of selected testing strategies in Massachusetts. Cumulative SARS-CoV-2 infections (Panels A-C) and mortality (Panels D-F) are plotted over time for early PCR-severe-only and two alternative testing strategies: Self-screen and PCR-all-repeat. Different starting dates for the implementation of testing strategies are shown (April 1, May 1, and June 1, 2020), with dash patterns indicating each start date, as listed in the figure key. Earlier implementation of the Self-screen strategy (orange lines) and the PCR-all-repeat testing with retesting strategy (green lines) correspond to lower cumulative infections over time. Panels A and D represent a slowing scenario in which the effective reproduction number (R_e_) on June 1, 2020 is 0.9. Panels B and E represent an intermediate scenario in which the R_e_ on June 1, 2020 is 1.3. Panels C and F represent a surging scenario in which the R_e_ on June 1, 2020 is 2.0. Abbreviations: R_e_, effective reproduction number; PCR, polymerase chain reaction

#### Cost-effectiveness

In one-way sensitivity analyses, the economically preferred strategy was most sensitive to test acceptance, the transmission reduction after a positive PCR test, and PCR test costs (Supplementary Tables 3-11). In the surging scenario, PCR-all-repeat would not be cost-effective if we assume low test acceptance (15%), half the transmission reduction after a positive test (33%), or double PCR test costs ($103). PCR-all-repeat would become cost-effective in the intermediate and slowing scenarios only with reductions in test costs (intermediate: ≤$13 slowing: ≤$3). If costs decrease for PCR assays, at many combinations of program and assay costs PCR-all-repeat strategy would be cost-effective (slowing and intermediate) or cost-saving (surging) (Supplementary Figure 4).

Holding other parameters equal to the base case, PCR-all-repeat would become cost-effective at an R_e_ value ≥1.8 (Supplementary Table 12). The frequency of repeat testing with PCR-all-repeat is also influential; in the surging scenario, PCR-all-repeat would no longer be cost-effective if tests occur more frequently than every 30 days (Supplementary Table 13). While total costs would vary widely with rates of COVID-19-like illness, cost-effectiveness conclusions would not change (Supplementary Table 14). Conclusions are robust to variations in estimates of life-years lost, or costs associated with lost productivity and averted COVID-related mortality (Supplementary Table 15 and 16).

## DISCUSSION

Using a microsimulation model, we projected the COVID-19 epidemic in Massachusetts from May 1, 2020 to November 1, 2020 under slowing, intermediate and surging epidemic growth scenarios, to examine the clinical and economic impact of five testing strategies.

Expanded PCR testing beyond those with severe symptoms would reduce morbidity and mortality across a range of epidemic scenarios. The response of the epidemic to “re-opening” is uncertain; in all R_e_ scenarios, we estimate substantial reductions in mortality (1.7-to 3.3-fold lower) with PCR-all-repeat compared to PCR-severe-only. Our R_e_ values encompass published estimates for MA during the study period [25–27]. Importantly, the slowing scenario likely reflects Massachusetts’s response through June 2020 [6], and the surging scenario provides important insight for elsewhere in the United States where infections are increasing.

We further estimate that if expanded PCR testing had been widely available in Massachusetts from April 1, 2020 to May 1, 2020, 106,200-184,200 infections and 80-100 deaths would have been averted during that one month alone. Given the time from infection to hospitalization and death (∼9 days and ∼28 days, respectively), earlier expanded testing might also have facilitated timely recognition of epidemic trends and closure policies. Policies that reduce R_e_ at scale (*e.g*., stay-at-home advisories), as occurred in Massachusetts even while PCR testing was scarce, are likely to be more effective than any of the modeled testing strategies [28,29]. Similar to conclusions from other studies [25,30–33], our findings suggest that looser restrictions on social distancing regulations (which can lead to a higher R_e_) would require more aggressive testing, paired with individual behavioral measures, to control the epidemic.

All the expanded screening strategies would lead to reductions in key hospital resource use as well as fewer days spent self-isolating compared to PCR-severe-only. In Massachusetts, an estimated 9,500 hospital beds and 1,500 ICU beds were available at the peak of the surge capacity, of which 3,800 and 1,440 were used [6,34]. None of the modeled scenarios exceeded peak hospital bed capacity even with PCR-severe-only; however, we projected 28-66% of available hospital beds would be needed by people with COVID-19. In all scenarios, we projected peak ICU bed use close to or exceeding capacity (1,200-3,600). While some assumptions are uncertain (*e.g*. proportion of people presenting to the hospital with severe disease, probability of ICU survival) the substantial burden of severe and critical illness we project in all scenarios has important implications for healthcare globally – resources redirected for COVID-related illness may jeopardize the ability to care for other diseases.

In all examined epidemic growth scenarios, PCR-any-symptom testing would be cost-saving compared to PCR-severe-only. To implement PCR-any-symptom, we estimate that 4,900-5,700 tests would be required daily. Even though PCR-all-repeat led to the least infections, mortality, and hospital resources used in all scenarios, it only would become cost-effective if the epidemic is surging or PCR assay cost is <10% base case values ($3 at R_e_ 0.9). At any R_e_ above 1.8, PCR-all-repeat would be the most efficient use of resources, unless test acceptance is very low (15%). Importantly, at these higher R_e_ values, screening the entire population only one time (PCR-all), would be an inefficient use without repeat screening for those testing negative (PCR-all-repeat).

In the slowing and intermediate scenarios, as of July 2020, Massachusetts would have test capacity to conduct the economically preferred strategy (estimated statewide tests conducted approximately 12,000/day) [6]. However, in the surging scenario, the projected average of 203,100 tests/day (36.6 million/180 days) required to conduct the cost-effective PCR-all-repeat strategy would greatly exceed current capacity. Large-scale testing has been achieved early in the epidemic in some settings: in March 2020, South Korea was testing 20,000 people/day [2]. Newer high throughput machines may process thousands of tests per day, rendering such an approach potentially feasible in the near future [35]. Additionally, the number of tests used for people without COVID-19 is uncertain; we thus assumed high rates of COVID-like-illness (adding approximately 2,800 tests/day) in the base case. However, it is likely, particularly in summer months, that fewer people would seek testing, reducing tests used by approximately 40%. Given that the economically preferred strategy changes depending on R_e_, implementation of the most cost-effective testing strategy will require careful planning and real-time epidemic monitoring in each setting to adapt to changing R_e_. While critical supply chain issues and other factors precluded widespread testing in the US early in the pandemic; even now, expanding testing capacity must remain a focus of national efforts.

The impact of any testing strategy depends on the actions that policymakers, employers, and individuals take in response. Our results emphasize how policies that support isolating people infected with COVID-19 are essential; when an individual is less adherent to self-isolation after a positive test (*i.e*., lower transmission reduction), the benefits of testing are greatly reduced. In Iceland, broad testing led to only 6% of the population being tested, with 34% of an invited random sample presenting for testing [1]. In the surging scenario, at low test acceptance rates (15%) among those with no or mild symptoms, PCR-all-repeat would no longer be cost-effective. In Massachusetts, SARS-CoV-2 testing does not require co-pays, and sufficient personal protective equipment permits safe testing [3,5]. Nevertheless, people may avoid testing due to concerns such as physical discomfort, missing work or stigma. While the Family Medical and Leave Act (FMLA) may provide support for those eligible who test positive (or if family members test positive), not all workers may be aware of their rights or have compliant employers [36]. Federal and setting-specific incentives for infected people to self-isolate should be considered (*e.g*., childcare or workplace incentives) [37].

This analysis has important limitations. First, we do not account for super-spreader transmission [38], and we assume homogenous population mixing; this may either over- or under-estimate the benefits of PCR testing. Second, we do not address supply chain lapses which could impact the feasibility of implementing these strategies. Third, we exclude several factors that would render testing even more cost-effective, including quality-of-life reductions due to COVID-related morbidity or self-quarantine-related mental health issues [39], school closure-related workforce gaps [40], and reductions in economic purchasing [31]. We also assume that transmissions vary with a constant daily rate by disease state; emerging data suggest that infectivity may be highest early after acquisition of the virus [41]. If true, testing strategies which diagnose people in early or asymptomatic stages of infection would be of higher value.

Testing people with any COVID-19-consistent symptoms would be cost-saving, compared to restricting testing to only those with symptoms severe enough to warrant hospitalization. Expanding SARS-CoV-2 PCR testing to asymptomatic people would reduce infections, deaths, and hospital resource use. When the COVID-19 pandemic is surging, further expansion to permit monthly re-testing after a negative test would be cost-effective.

## Data Availability

The analysis was conducted from the data presenting in the manuscript and supplemental appendix.

## FUNDING

This work was supported by the Eunice Kennedy Shriver National Institute for Child Health and Human Development [K08 HD094638 to AMN], the National Institute of Allergy and Infectious Disease at the National Institutes of Health [T32 AI007433 to AM], and the Wellcome Trust [210479/Z/18/Z to GH].

The content is solely the responsibility of the authors, and the study’s findings and conclusions do not necessarily represent the official views of the National Institutes of Health, the Wellcome Trust, or other funders.

## ACKNOWLEDGMENTS

The authors gratefully acknowledge Christopher Alba, Giulia Park, and Tijana Stanic for their assistance in preparing the manuscript for publication.

## AUTHOR ROLES

All authors contributed substantively to this manuscript in the following ways: study and model design (all authors), data analysis (AMN, ACB), interpretation of results (all authors), drafting the manuscript (AMN, ACB, AM, PK), and critical revision of the manuscript (all authors) and final approval of submitted version (all authors).

## CONFLICTS OF INTEREST AND FINANCIAL DISCLOSURES

The authors have no conflicts of interest or financial disclosures.

## Notes

### Competing Interest Statement

The authors have declared no competing interest.

### Author Declarations

Approved by the Partners Human Research Committee under Protocol 2020P000967 All necessary patient/participant consent has been obtained and the appropriate institutional forms have been archived.

